# Assessment of The Relationship of REMS and MEWS Scores with Prognosis in Patients Diagnosed with Covid-19 Admitted to the Emergency Department

**DOI:** 10.1101/2021.01.24.21250384

**Authors:** Behlul Bas, Mucahit Senturk, Tugce Nur Burnaz, Kubilay Timur, Asim Kalkan

**Author notes:** **Correspondence Address**: Dr. Behlul Bas, Okmeydani Training and Research Hospital, Department of Emergency Medicine, İstanbul / TURKEY, **E-mail**. **DECLARATIONS**. **Author contributions**: B.B., M.S., T.N.B., K.T., and A.K. contributed in concept; T.N.B, M.S. and A.K helped in design; B.B, M.S., A.K. helped in supervision; M.S., T.N.B., K.T., A.K. helped in resources; B.B., M.S., T.N.B., K.T. contributed to materials; B.B., M.S., T.N.B., K.T., and A.K. contributed to data collection and/or processing; B.B., M.S., T.N.B., K.T., and A.K. helped in literature search; B.B., M.S., T.N.B., K.T., and A.K. helped in writing the manuscript; and B.B., M.S., T.N.B., K.T., and A.K. helped in critical review.

## Abstract

**Aim:** With the rapid and global increase in COVID-19 cases, it is becoming important to identify patients with a risk of mortality and patients that need hospitalization. The aim of this study is to try to predict the mortality rate of COVID patients admitted to the emergency department with rapid scoring systems such as REMS and MEWS and their clinical termination in the emergency department at the end of the first month.

**Method:** We have designed this study to be a single-centered, prospective and an observational study. A total of 392 patients diagnosed with COVID-19, who were admitted to the emergency department in a 1-month period, were included in the study. REMS and MEWS scores were calculated for each case. Demographic data of patients, clinical outcomes such as discharge, service hospitalization, ICU hospitalization, and first-month mortality were analysed based on these scores. ROC curves were analysed to determine the cut-off value with the help of which REMS and MEWS scores can predict 1-month mortality and hospitalization.

**Results:** Out of the 392 patients included in the study, the 43.4% (n=170) were female and 56.6% (n=222) were male. The average age of our patients was 48.98±19.49 years. The 1-month mortality rate of our patients was 4.3% (n=17). At the end of the first month, the mortality of patients with a comorbid disease was higher than those who did not (p<0.01). The average of the REMS score was higher in patients with an average mortality of (7.24±3.77) than in patients without it (2.87±3.09), and there was a statistically significant difference between them (p<0.01). Similarly, the average of the MEWS score was higher in patients with an average mortality of (2.76±1.86) than in patients without it (1.65±1.35), and there was a statistically significant difference (p<0.01). The REMS score of patients admitted to the service was higher than that of patients discharged (p<0.01). When the REMS score was determined as 3 cut-off value in ROC analysis, service hospitalization was 5 times higher in patients with a REMS score of 3 and above than in those who were discharged (OR: 1:5.022 95% CI: 3.088-8.168)). REMS and MEWS scores were also higher in ICU patients than in discharged patients (p<0.01).

**Conclusion:** In predicting the 1-month mortality of ER patients diagnosed with COVID-19, REMS and MEWS scoring systems can be useful and guiding in determining the patients who need hospitalization for emergency physicians. The use of these scoring systems in emergency departments can help predict the clinical outcomes of patients at the time of the initial evaluation, and can also be a practical method of predicting the prognosis of the patients.

## Introduction

Coronaviruses are RNA viruses that cause diseases by affecting multiple systems in humans and other living things [1]. Up until the last few years, 6 types of coronaviruses that caused the disease in humans had been known. A new type of coronavirus was discovered in Wuhan, China, in the last months of 2019, after an increase in pneumonia cases with an unknown factor. This virus was named COVID-19. Some of the patients infected with the virus were asymptomatic, and some were admitted to the hospital with symptoms such as fever, cough, weakness, runny nose, chest pain, diarrhea and respiratory failure [2,3].

The clinical course of patients suffering from COVID-19 infection was linked to several risk factors such as age, gender, presence of comorbid disease and smoking history in some studies [4-7]. Many scoring systems have also been investigated in these patient groups in order to predict the clinical course and the course of patients after hospitalization. To date, the use of scores that predict early mortality in emergency departments has been a rational approach, as it ensures close follow-up and treatment of patients. Examples of these scorings are REMS and MEWS scoring [8]. Research on early warning scores that can predict prognosis during emergency department admission in COVID-19 infection is limited. One of these studies has been conducted only to predict the mortality of patients in intensive care, and the other has been conducted to predict 48-hour and 7-day mortality with some scoring systems [9-10].

The aim of this study is to evaluate the availability of REMS and MEWS scores to predict 1-month mortality and emergency department clinical outcome of patients with COVID-19 infection admitted to the emergency department.

## Methods

### Study Design

This study has been designed as a single-centered, prospective and an observational study. Patients diagnosed with COVID-19 who were admitted to XXX Hospital Emergency Medicine Clinic were included in our study. Our hospital emergency department is a tertiary, multidisciplinary hospital where 500.000-550.000 patients are cared for annually. Our study was completed within 1 month on patients diagnosed with COVID-19 out of patients admitted to the emergency department pandemic area of our hospital. Our hospital has about 400 patients applying daily with the suspicion of COVID-19 to the emergency department pandemic area, and about 40 patients are diagnosed with COVID-19. Written informed consent was obtained from the patients for their anonymized information to be published in this article.

### Study Subjects and Settings

Patients diagnosed with COVID-19 and admitted to our hospital’s Emergency Medicine Clinic between 07/06/2020 and 07/07/2020 were included in this study. All patients over the age of 18 who were clinically, radiologically or laboratory diagnosed with COVID-19 and who gave consent to participate in the study were included in our study. Pregnant patients, patients under 18, patients who did not consent to participate in the study, patients who had suffered trauma, pregnant women, and patients on drugs that are primarily effective on the central nervous system such as antidepressants, antipsychotics, etc. were not included in the study. The local Ethics Committee approved the study. The study was also in line with the Declaration of Helsinki.

### Data Analysis and Measurement

Demographic data of the patients included in the study, their symptoms during admission, and information about their clinical outcome in the emergency department, such as hospitalization or discharge, have been collected and analysed. This data contained all the information needed to calculate REMS and MEWS scores. REMS to calculate the score; the state of consciousness, GKS, Average blood pressure (mmHg) heart rate (beats/minute) respiratory rate (breaths/minute), fever (°C), Partial oxygen saturation (%) and patient age (years) information to calculate MEWS score; state of consciousness, systolic blood pressure (mmHg), heart rate (beats/min), body temperature (°C), respiratory rate (breath/min) data were used. REMS and MEWS scores calculated based on these data have also been recorded. The 1-month mortality information of patients enrolled in the study has been examined and analyzed through hospital data.

### Statistical Analysis

The NCSS (Number Cruncher Statistical System) (Kaysville, Utah, USA) program was used for statistical analysis. Descriptive statistical methods (mean, standard deviation, median, frequency, ratio, minimum, maximum) were used while evaluating the study data. The suitability of quantitative data for normal distribution was tested with the Kolmogorov-Smirnov, Shapiro-Wilk test and graphical evaluations. Student’s t Test was used in two-group comparisons of quantitative data with a normal distribution, and the Mann Whitney U test was used in two-group comparisons of data without a normal distribution. Kruskal Wallis test and Bonferroni Dunn test were used for binary comparisons of three and above groups that did not show Normal distribution. Pearson Chi-Square test, Fisher-Freeman-Halton Exact test and Fisher’s Exact test were used to compare qualitative data. Diagnostic screening tests (sensitivity, specificity, PKD, NKD) and ROC Curve analyses were used to determine cut off for parameters. Significance was evaluated at a level of at least p<0.05.

## Results

Our study included 455 patients. A total of 63 patients whose data was recorded incomplete and who were using antidepressants were excluded from the study. Of the 392 patients included in our study, 43.4% (n=170) were female and 56.6% (n=222) were male. The average age of our patients was 48.98±19.49 years. The age of the cases with mortality at the end of the 1st month was higher (p<0.01). But no statistically significant differences have been found between gender distributions and 1-month mortality (p>0.05). The most common additional disease of our patients was hypertension, followed by diabetes mellitus and ischemic heart disease. The most common symptoms were shortness of breath and cough. These symptoms were followed by fever, headache, and myalgia. Information about our patient’s vital signs, average REMS and MEWS scores, states of consciousness, comorbid diseases, symptoms and mortality rates are indicated in Table-1 (Table-1).

The mortality rate of our patients at the end of the first month was 4.3% (n=17). At the end of the first month, the mortality of patients with a comorbid disease - was 20.8 times higher than those without a comorbid disease, and a statistically significant difference was found between them (p<0.01) ((OR: 1:20.810 (95% CI: 2.731-158.539). The average REMS score was higher in patients who died (non-survival) (7.24±3.77) than in patients who survived (2.87±3.09), and there was a statistically significant difference between them (p<0.01). The average MEWS score was also higher in non-survival patients (2.76±1.86) than in survivor patients (1.65±1.35), and there was a statistically significant difference between them (p<0.01). In other words, at the end of the first month of the disease, REMS and MEWS scores were higher in cases with mortality (Table 2). Based on this significance, calculating the cut off point for REMS and MEWS scores was taken into consideration.

**Table 1:**
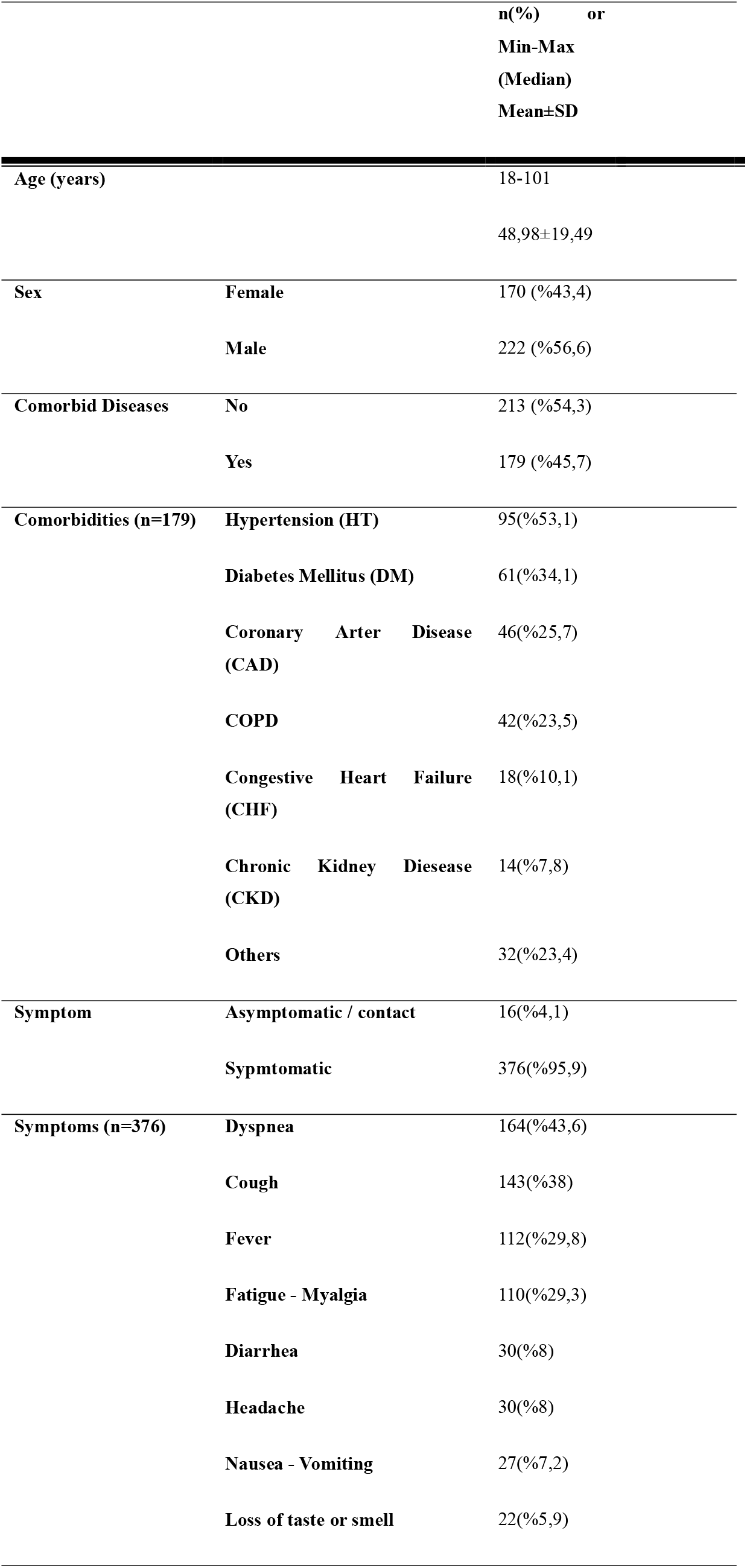
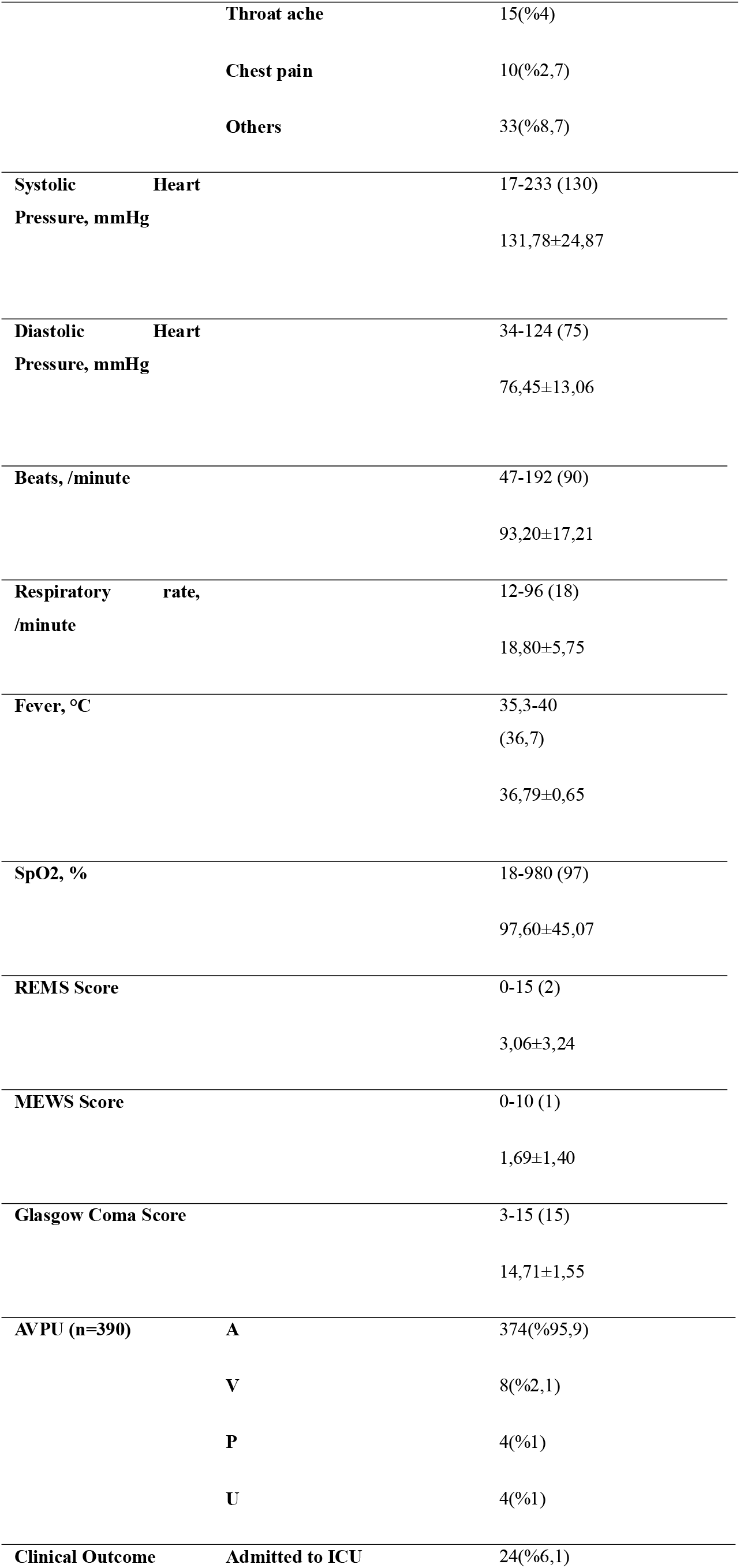
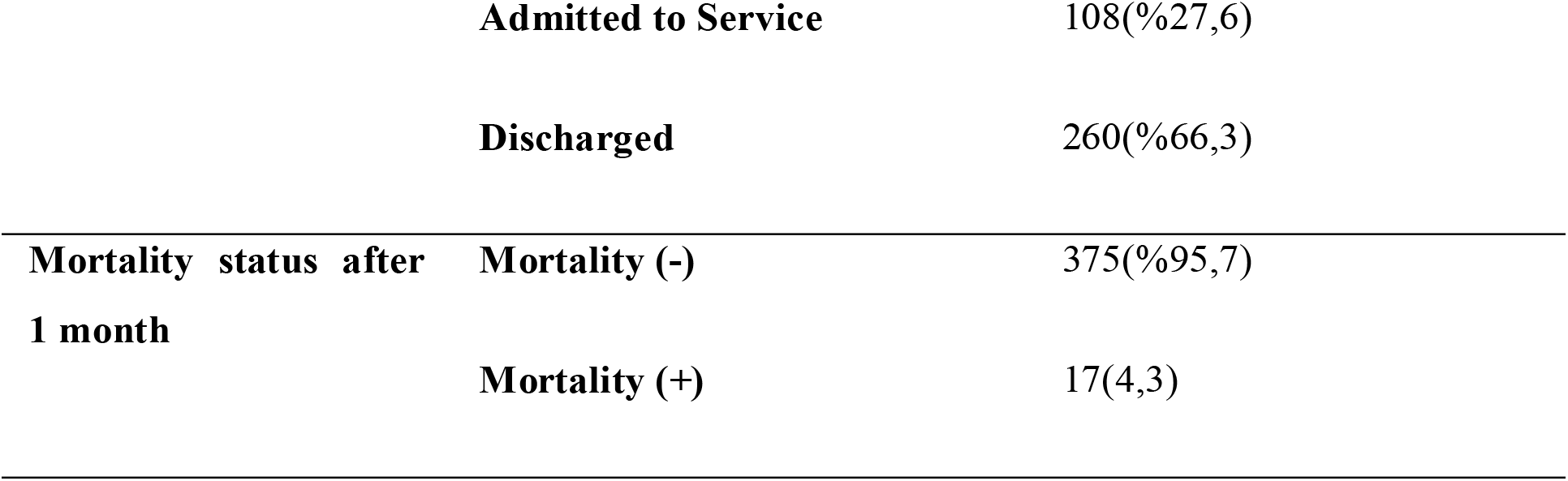
The Distribution Of Demographic Characteristics

**Table 2:** The Evaluation of Mortality Results After 1 Month According to Demographic Characteristics and Comorbid Diseases

|  |  | **Mortality status after 1 month** |  |  |
| --- | --- | --- | --- | --- |
|  |  | **No (n=375)** | **Yes (n=17)** |  |
|  |  | **n (%)** | **n (%)** | **p** |
| **Age (years)** | *Min-Max (Median)* | 10-101 (48) | 45-88 (80) | <sup>a</sup>**0,001\*\*** |
|  | *Mean±SD* | 47,83±18,98 | 73,76±15,01 |  |
| **Sex** | **Female** | 164 (43,7) | 6 (35,3) | <sup>b</sup>**0,492** |
|  | **Male** | 211 (56,3) | 11 (64,7) |  |
| **Comorbid Disease** | **No** | 212 (56,5) | 1 (5,9) | <sup>b</sup>**0,001\*\*** |
|  | **Yes** | 163 (43,5) | 16 (94,1) |  |
| **REMS Score** | *Min-Max (Median)* | 0-15 (2) | 0-14 (7) | <sup>d</sup>**0,001\*\*** |
|  | *Mean±SD* | 2,87±3,09 | 7,24±3,77 |  |
| **MEWS Score** | *Min-Max (Median)* | 0-10 (1) | 0-7 (3) | <sup>d</sup>**0,007\*\*** |
|  | *Mean±SD* | 1,65±1,35 | 2,76±1,86 |  |

The cut-off point of groups for REMS score has been determined as 5 and above. When 1-month mortality for 5 cut-off values of REMS score was examined; sensitivity was found to be 82.35%, specificity 71.47%, positive estimation 11.57%, negative estimation 98.89% and accuracy was 71.94%. In the ROC curve obtained (Figure 1), AUC 81.8% standard error was determined as 5.3%. A statistically significant association was found between mortality at the end of the first month and the 5 cut-off values of the REMS score (p<0.01), and the risk of mortality was found to be 11.7 times higher in patients with REMS score 5 and above (OR: 11,688 (95% CI: 3.293-41,493)). For the MEWS score, the cut-off point was 3 and higher, and for this estimated value; sensitivity was 52.94%, specificity was 82.40%, positive estimate was 12.0%, negative estimate was 97.48%, and accuracy was 81.12%. In the obtained ROC curve, AUC 68.1% standard error was determined as 7.5%. At the end of the first month, a statistically significant relationship was found between the mortality rate and the 3 cut-off values of the MEWS score (p<0.01). In patients with a MEWS score of 3 and above, the risk of mortality at the end of the first month is 5.3 times higher (OR: 1:5.267 (95% CI: 1.960-14.157)) (Table 4).

**Figure 1:**
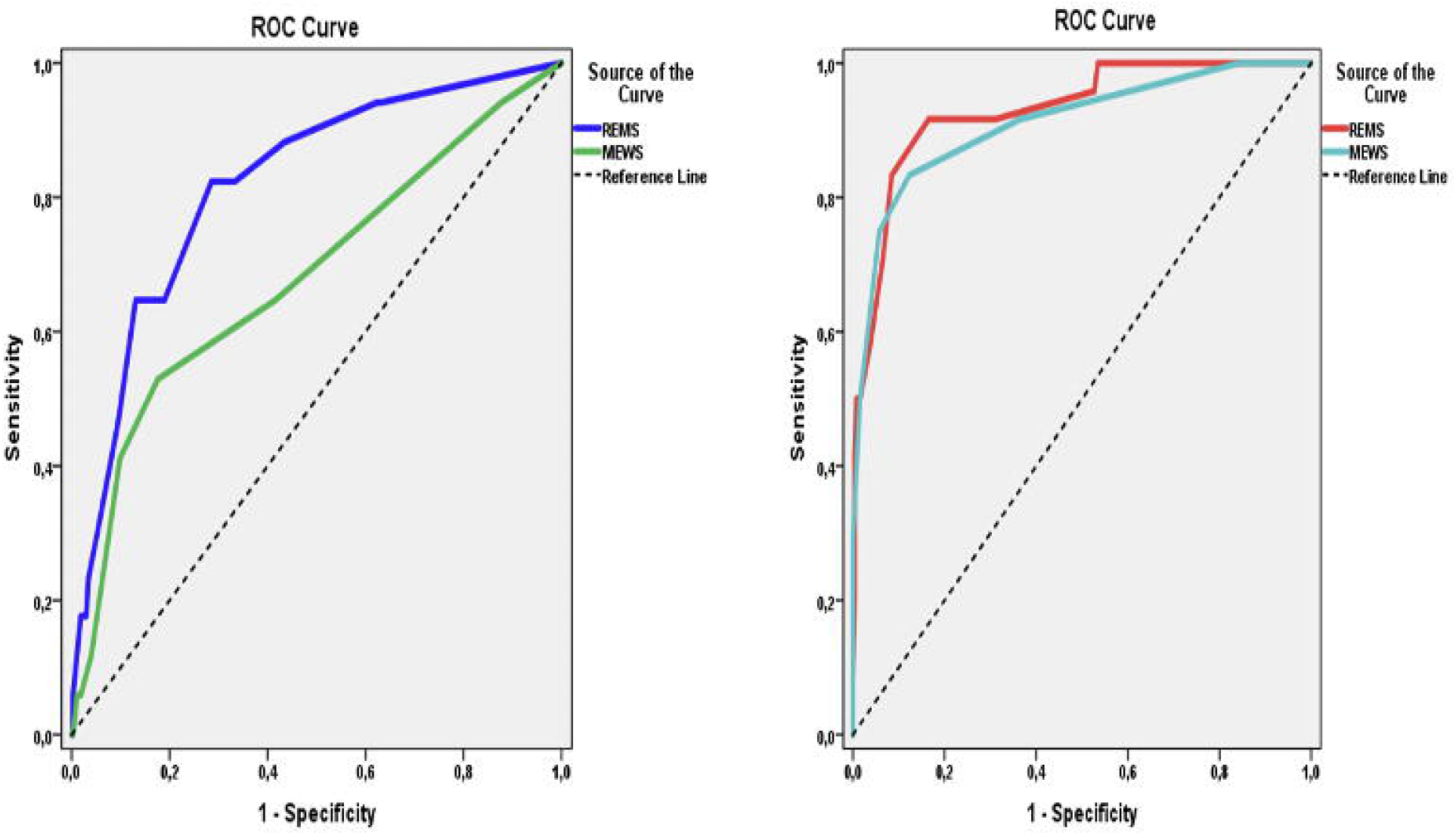
ROC curve for REMS and MEWS scores based on mortality (left) and ICU hospitalization and discharge (right) after 1 month

In the study of clinical outcome of the ER patients, the incidence of comorbid disease was higher in ICU and hospitalized patients than in discharged patients (p=0.001; p=0.001 p<0.01, respectively). The most common comorbid disease in ICU patients was hypertension, which was followed by DM, IKH and COPD. REMS scores of ICU and service hospitalization cases are higher than those of discharge cases (p<0.01). Similarly, MEWS scores in ICU and service hospitalization were higher than in discharged cases (p=0.001; p=0.031; p<0.05, respectively). In addition, REMs and MEWS scores of patients admitted to ICU were also higher than patients admitted to the service (p<0.01).

A statistically significant relationship was found between ICU admission and the discharge status and the 5 cut-off values of the REMS score (p<0.01). In patients with a REMS score of 5 and above, the risk of ICU hospitalization is 55.5 times higher than in those who have been discharged (OR: 1:55.512 95% CI: 12.586-244,847)) (Table 4).

According to ICU hospitalization and discharge groups, the cut off point for MEWS score was determined as 3 and higher. For 3 cutting values of the mews score, sensitivity is 83.33%, specificity is 87.69%, positive estimation is 38.46%, negative estimation is 98.28% and accuracy is 87.32%. In the obtained ROC curve, the underlying area was found to be 91.1% standard error 3.6% (Figure 1). A statistically significant relationship has been found between ICU admission/discharge status and the 3 cut-off values of the MEWS score (p=0.001; p<0.01). In patients with a MEWS score of 3 and above, the risk of ICU hospitalization is 35.63 times higher than in those who have been discharged (OR: 1:35.625 (95% CI: 11ch445-110.890)) (Table 5).

REMS and MEWS scores were found to be higher in patients with service hospitalization than in patients who were discharged (p<0.01, p<0.05) (Table 4). Based on this significance, the calculation of the cut off point for REMS and MEWS scores was taken into consideration. The cut-off point for REMS score was determined as 3 and higher for groups who were hospitalized and discharged. For 3 cut-off values of REMS score, sensitivity is 69.44%, specificity is 68.85%, positive estimation is 48.08%, negative estimation is 84.43% and accuracy is 69.02%. In the obtained ROC curve, the underlying area was determined as 73.7% standard error 2.9% (Figure 2). In patients with a REMS score of 3 and above, service admission was 5.022 times higher than in those discharged (OR 1:5.022 (95% CI: 3.088-8.168)) (Table 5). Cut off point for MEWS score was determined as 1 and above in the same groups. For the 1 cut-off value of the MEWS score, sensitivity is 94.44%, specificity is 15.77%, positive estimation is 31.78%, negative estimation is 87.23% and accuracy is 38.86%. In the resulting ROC curve, the underlying area was found to be 58.4% and standard error was found to be 3.2% (Figure 2) a statistically significant relationship has also been found between service hospitalization and discharge status and the mews score cut-off value of 1 (p<0.01). In cases with a MEWS score of 1 and above, service hospitalization is 3.183 times higher than in those discharged (OR: 1: 3.183 (95% CI: 1.309-7.737)) (Table 5). REMs and MEWS scores were found to be higher in ICU hospitalization cases than in service hospitalization cases (p<0.01) (Table 3). Based on this significance, it was considered to calculate the cut-off point for REMS and MEWS scores. The cut-off point for REMS score was determined as 7 and above for ICU hospitalization and service hospitalization groups. For 7 cut-off values of REMS score, sensitivity is 70.83%, specificity is 75.93%, positive estimation is 39.53%, negative estimation is 92.13% and accuracy is 75.0%. In the obtained ROC curve, the underlying area was found to be 81.5% standard error 5.1% (Figure 2). A statistically significant relationship was found between ICU hospitalization, service hospitalization and the 7 cut-off values of the REMS score (p<0.01). In patients with a REMS score of 7 and above, the risk of ICU hospitalization is 7.66 times higher than in those with a service hospitalization (OR: 1:7.659 (95% CI: 2.862-20.501)) (Table 5).

**Table 3:** Evaluations According To The Patient’s Clinical Outcome

|  |  | Patient's Clinical Outcome |  |  | *p* |
| --- | --- | --- | --- | --- | --- |
|  |  | Admitted to ICU (n=24) | Admitted to Service (n=108) | Discharged (n=260) |  |
|  |  | n (%) | n (%) | n (%) |  |
| **Comorbid Diseases** | **No** | 3 (12,5) | 34 (31,5) | 176 (67,7) | <sup>b</sup>0,001** |
|  | **Yes** | 21 (87,5) | 74 (68,5) | 84 (32,3) |  |
| **Comorbidities** | **HT** | 14 (58,3) | 39 (36,1) | 42 (16,2) | <sup>b</sup>0,001** |
|  | **DM** | 7 (29,2) | 28 (25,9) | 26 (10,0) | <sup>b</sup>0,001** |
|  | **COPD** | 5 (20,8) | 15 (13,9) | 22 (8,5) | <sup>b</sup>0,079 |
|  | **CAD** | 8 (33,3) | 22 (20,4) | 16 (6,2) | <sup>b</sup>0,001** |
|  | **CKD** | 3 (12,5) | 6 (5,6) | 5 (1,9) | <sup>e</sup>0,015* |
|  | **CHF** | 4 (16,7) | 10 (9,3) | 4 (1,5) | <sup>e</sup>0,001** |
|  | **Malignancy** | 2 (8,3) | 5 (4,6) | 7 (2,7) | <sup>e</sup>0,203 |
|  | **CVO** | 5 (25,0) | 5 (4,6) | 0 (0) | <sup>e</sup>0,001** |
|  | **Others** | 2 (8,3) | 7 (6,5) | 8 (3,1) | <sup>e</sup>0,145 |
| **REMS Score** | *Min-Max (Medyan)* | 1-15 (9) | 0-12 (5) | 0-13 (2) | <sup>f</sup>0,001** |
|  | *Mean±SD* | 8,88±3,77 | 4,40±3,00 | 1,97±2,41 |  |
| **MEWS Score** | *Min-Mak (Medyan)* | 1-10 (4,5) | 0-6 (1) | 0-5 (1) | <sup>f</sup>0,001** |
|  | *Mean±SD* | 4,63±2,12 | 1,74±1,13 | 1,40±1,06 |  |

<sup>b</sup>Pearson chi-square Test <sup>e</sup>Fisher Freeman Halton Exact Test <sup>f</sup>Kruskal Wallis Test
\*\**p*<0.01 \**p*<0.05

**Table 4:**
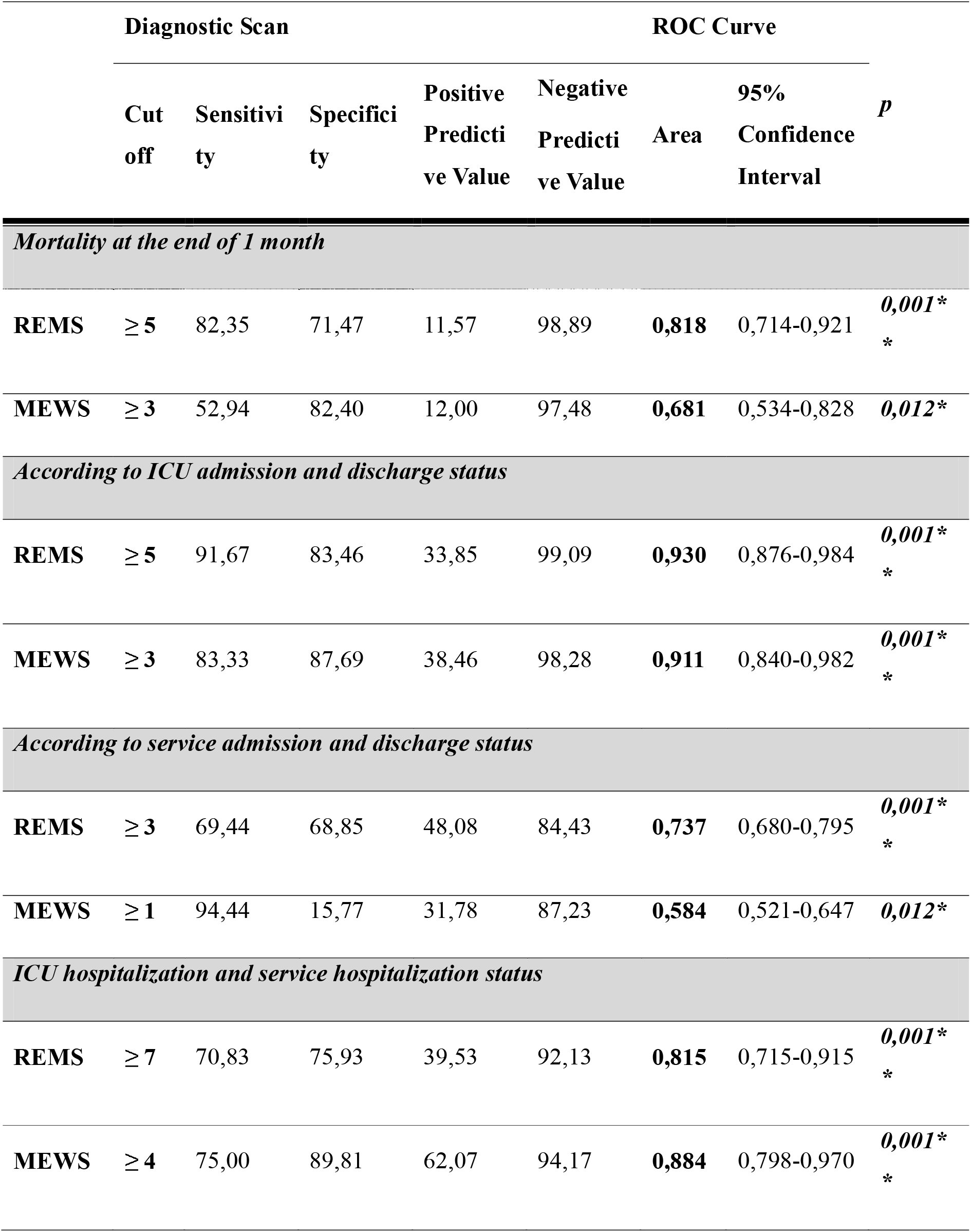
Diagnostic Tests and ROC Curve Results for REMS and MEWS Scores

**Table 5:** Relationship between REMS and MEWS scores (Cut-off values)

|  | REMS < 5 | REMS ≥ 5 | P | MEWS < 3 | MEWS ≥ 3 | P |
| --- | --- | --- | --- | --- | --- | --- |
| **Mortality (-)** | 268 (71,5) | 107 (28,5) | <sup>b</sup>0,001** | 309 (82,4) | 66 (17,6) | <sup>c</sup>0,001** |
| **Mortality (+)** | 3 (17,6) | 14 (82,4) |  | 8 (47,1) | 9 (52,9) |  |
|  | REMS < 5 | REMS ≥ 5 |  | MEWS < 3 | MEWS ≥ 3 |  |
| **Discharged** | 217 (83,5) | 43 (16,5) | <sup>b</sup>0,001** | 228 (87,7) | 32 (12,3) | <sup>c</sup>0,001** |
| **Admitted to ICU** | 2 (8,3) | 22 (91,7) |  | 4 (16,7) | 20 (83,3) |  |
|  | REMS < 3 | REMS ≥ 3 |  | MEWS < 1 | MEWS ≥ 1 |  |
| **Discharged** | 179 (68,8) | 81 (31,2) | <sup>b</sup>0,001** | 41 (15,8) | 219 (84,2) | <sup>b</sup>0,008** |
| **Admitted to Service** | 33 (30,6) | 75 (69,4) |  | 6 (5,6) | 102 (94,4) |  |
|  | REMS < 7 | REMS ≥ 7 |  | MEWS < 4 | MEWS ≥ 4 |  |
| **Admitted to Service** | 82 (75,9) | 26 (24,1) | <sup>b</sup>0,001** | 97 (89,8) | 11 (10,2) | <sup>b</sup>0,001** |
| **Admitted to ICU** | 7 (29,2) | 17 (70,8) |  | 6 (25) | 18 (75) |  |

<sup>b</sup>Pearson chi-square Test
<sup>c</sup>Fisher's Exact Test
\*\*p<0.01

**Figure 2:**
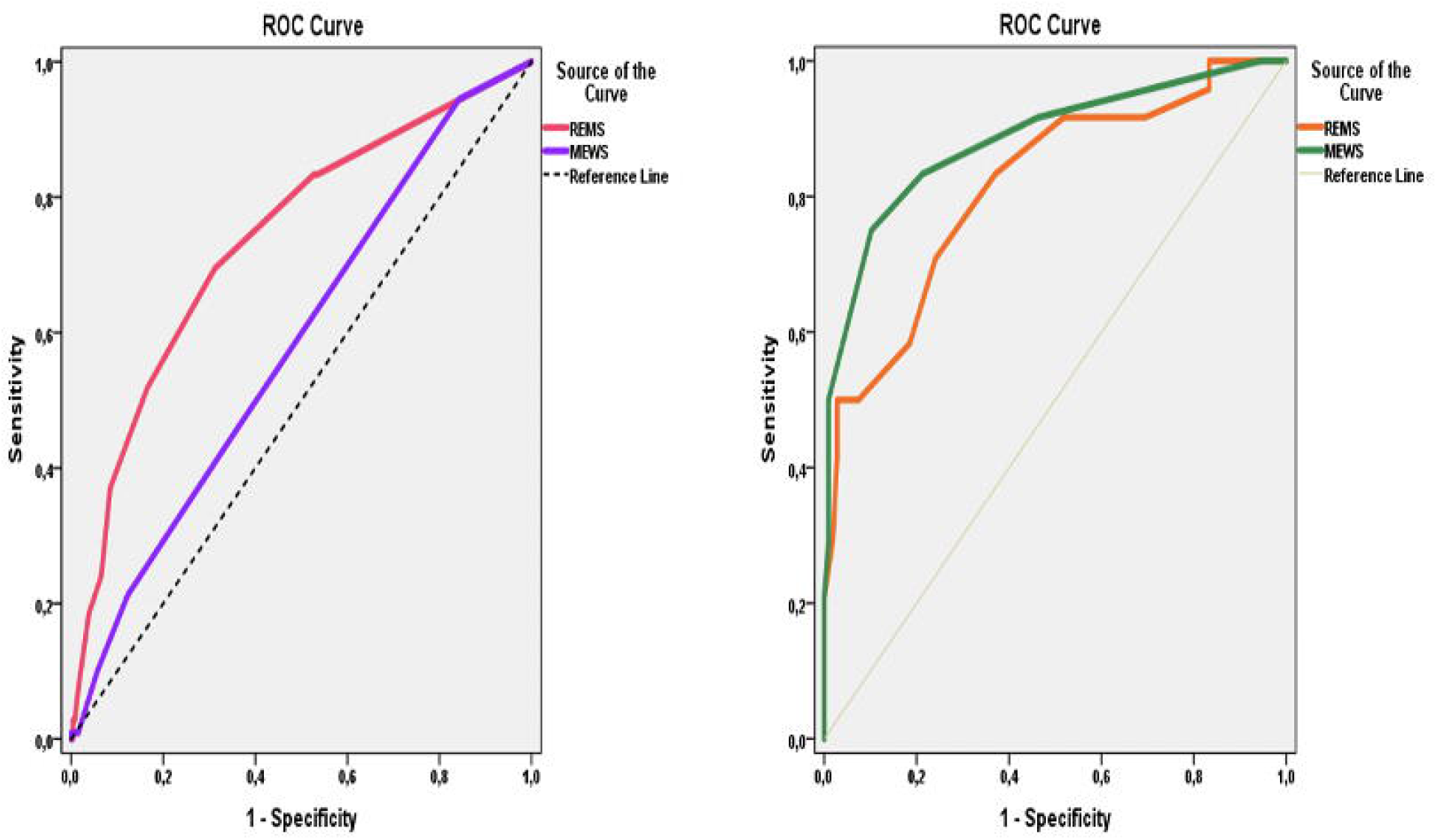
ROC curve for REMS and MEWS scores according to service admission or discharge (left) and ICU hospitalization or service hospitalization (right)

The cut-off point for MEWS score for ICU hospitalization and service hospitalization groups was determined as 4 and above. Sensitivity is 75.00%, specificity is 89.81%, positive estimation is 62.07%, negative estimation is 94.17% and accuracy is 87.12% for 4 cut-off values of the MEWS score. In the resulting ROC curve, the underlying area has been found to be 88.4% and standard error to be 4.4% (Figure 2). A statistically significant relationship has been found between ICU hospitalization, service hospitalization and the 4 cut-off values of the MEWS score (p=0.001; p<0.01). In patients with a MEWS score of 4 and above, the risk of ICU hospitalization is 26.46 times higher than in those with a service hospitalization (OR: 1:26.455 (95% CI: 8.678-80.648)) (Table 5).

## Discussion

Our study has shown that there is no special rapid scoring system used to predict the prognosis of COVID-19. In this study, we have tried to determine the prognosis of these patients using REMS and MEWS scoring systems. In our study, we have found that COVID-19 patients with high REMS and MEWS scores had higher hospitalization, intensive care unit admission and 1-month mortality rate. In addition, both mortality and hospitalization rates of patients with comorbidity were higher. In previous studies, it was emphasized that comorbidity is important for hospitalization and admission to intensive care [4,11,12]. Similar to these studies, patients with comorbidity had higher mortality at the end of the first month in our study, too.

MEWS is a modified version [13] of the Early Warning Score (MEWS) developed by Subbe et al in 2000. As an easily computable per-patient tool in a busy clinical area, MEWS can help identify the need for early intervention while evaluating emergency patients. Moreover, MEWS is a scoring system which uses vital parameters and which is calculated by systolic blood pressure, heart rate, respiratory rate, body temperature and AVPU scale [14]. On the other hand, REMS is a scoring system developed by Olsson et al, and which uses Glasgow Coma Scale instead of AVPU unlike Mews; the age of the patient is also included in the scoring while using REMS [15]. The main reason we have received high scores in these two scoring systems is the fact that the score points increase as the vital signs deteriorate. The study of Hu et al supports the findings of our study. Hu et al used the REMS and MEWS rapid scoring systems, which are normally used in the emergency department, on Covid-19 patients in critical condition. Their study included 105 patients and they noted that mortality would be high at certain cut-off values in the REMS score and MEWS score. In light of these data, they argued that the REMS scoring system is better than MEWS in predicting mortality in critical patients [10, 16,17]. In our study, we tried to predict mortality and prognosis in all COVID-19 patients admitted to the emergency department using these two scoring systems.

The advantage of our study compared to other studies is that it has examined all applications and reviewed service hospitalization, intensive care hospitalization and mortality altogether. Hu et al evaluated only mortality for each cut off value in critical patients in intensive care [10]. In our study, it has been determined that the patient’s prognosis can be defined based on the cut off values obtained.

When we compared REMS and MEWS scores in our study, we found that the REMS score of 5 points and above was superior to the MEWS score of 3 points and above. A positive value as AUC value 0.818 was determined for REMS score 5 points and above. Although the high REMS score was not very strong in determining mortality (PPV=11.57) it was very successful in determining that there would be no mortality of patients below 5 points (NPV=98.89). We can attribute this to the fact that the number of patients we lost was only 17. A MEWS score of 3 or higher is also a useful method for determining patient mortality, but it is not as strong as REMS (AUC=0.681)

According to another finding of our study, REMS is again found to be superior than the MEWS score to distinguish when the patients will be admitted to intensive care or when they will be discharged. Patients with a REMS score of 5 points and above are more likely to be admitted to intensive care. (AUC = 0.930). This value was as strong as REMS score for 3 points and above values for the MEWS score (AUC=0.911). Therefore, it should be noted that patients above 5 and 3 points in REMS and MEWS scores, respectively, are more likely to be admitted to intensive care.

Also another substantial discovery we made was that the MEWS score slightly exceeded the REMS score when patients admitted to the service and admitted to the intensive care unit were compared. Patients with a MEWS score of 4 points and above, and patients with a REMS score of 7 points and above, were mostly admitted to intensive care. That is why we believe that the MEWS score is a good scoring system that will be used in the emergency department in order to be admitted to the ICU.

### Limitation

The most important limitation of our study is that our number of patients and mortality rate are low. However, the study that has the largest number of patients in the literature happens to be our study. Our second limitation is that in our study, we did not separate the age groups of our patients based on decades. If we grouped patients based on their ages, we would probably find different mortality scores depending on the age group. Because in the REMS scoring system, different scores are received from different age groups. Another limitation is that we have not determined the mortality based on the treatment given to our patients. Keeping this factor in mind for other prospective studies on this subject will contribute to the literature.

## Conclusion

To sum up, our study has shown that REMS and MEWS scoring systems can be useful and guiding for emergency physicians in determining the 1-month mortality of COVID-19 diagnosed patients and in determining which patients need to be hospitalized. The use of these scoring systems in emergency departments can help predict the clinical endings of patients in the initial evaluation, and can also be a practical method of predicting the prognosis of patients.

## Data Availability

Data obtained from the patients who attend Okmeydani Training and Research Hospital, Emergency Department.

## Acknowledgements

The authors thank the dispatch and paramedics in the study area.

## References

1. Zhu N, Zhang D, Wang W, et al. A novel coronavirus from patients with pneumonia in China, 2019. N Engl J Med. 2020;382:727–33.

2. Corman VM, Lienau J, Witzenrath M. Coronaviruses as the cause of respiratory infections. Internist. 2019;60:1136–45.

3. Hoffmann M, Kleine-Weber H, Schroeder S, et al. SARS-CoV-2 Cell Entry Depends on ACE2 and TMPRSS2 and Is Blocked by a Clinically Proven Protease Inhibitor. Cell. 2020;181:271-80.e8.

4. Chen N, Zhou M, Dong X, et al. Epidemiological and clinical characteristics of 99 cases of 2019 novel coronavirus pneumonia in Wuhan, China: a descriptive study. Lancet. 2020;395:507–13.

5. Sakurai A, Sasaki T, Kato S, et al. Natural History of Asymptomatic SARS-CoV-2 Infection. N Engl J Med 2020; 383:885–6

6. Bajema KL, Oster AM, McGovern OL, et al. Persons Evaluated for 2019 Novel Coronavirus - United States, January 2020. MMWR Morb Mortal Wkly Rep. 2020;69:166– 70.

7. Chan JFW, Yuan S, Kok KH, et al. A familial cluster of pneumonia associated with the 2019 novel coronavirus indicating person-to-person transmission: a study of a family cluster. Lancet. 2020;395:514–23.

8. Smith GB, Prytherch DR, Meredith P, Schmidt PE, Featherstone PI. The ability of the National Early Warning Score (NEWS) to discriminate patients at risk of early cardiac arrest, unanticipated intensive care unit admission, and death. Resuscitation. 2013;84:465– 70.

9. Ciceri F, Castagna A, Rovere-Querini P, et al. Early predictors of clinical outcomes of COVID-19 outbreak in Milan, Italy. Clin Immunol. 2020;217:108509

10. Hu H, Yao N, Qiu Y. Comparing Rapid Scoring Systems in Mortality Prediction of Critically Ill Patients With Novel Coronavirus Disease. Acad Emerg Med. 2020;27:461– 68

11. Chen Z, Cheng Z, Zhang X, et al. Clinical manifestations and CT characteristics sa corona virus disease 2019 (COVID-19). Radiol Pract. 2020; 3:286–90.

12. Zhu YC, Tan L, Liu L, Li KZ, Qi WY, Hu X. Comparative analysis of characteristics and medications between corona virus disease 2019 and several acute reproductive syndrome. Clin Med J. 2020; 18:15–23.

13. Covino M, Sandroni C, Santoro M, Sabia L, Simeoni B, Bocci MG, Ojetti V, Candelli M, Antonelli M, Gasbarrini A, Franceschi F. Predicting intensive care unit admission and death for COVID-19 patients in the emergency department using early warning scores. Resuscitation. 2020;9;156:84-91.

14. Subbe CP, Kruger M, Ruther FB. Validation of a modified early warning score in medical admissions. QJ Med. 2001;94:521–6.12

15. Olsson T, Terent A, Lind L. Rapid Emergency Medicine Score can predict long-term mortality in nonsurgical emergency department patients. Acad Emerg Med. 2004;11:1008–13.

16. Montenegro SM, Rodrigues CH. Evaluation of the performance of the modified early warning score in a Brazilian public hospital. Rev Bras Enferm 2019;72:1428–34.

17. Olino L, Gonçalves AC, Strada JK, et al. Effective communication for patient safety: transfer note and Modified Early Warning Score. Rev Gaucha Enferm. 2019;40:1–8

